# Adaptive Cardiac Resynchronization Therapy Effect on Electrical Dyssynchrony (aCRT-ELSYNC): a randomized controlled trial

**DOI:** 10.1101/2020.09.14.20194415

**Authors:** Kazi T. Haq, Nichole M. Rogovoy, Jason A. Thomas, Christopher Hamilton, Katherine J. Lutz, Ashley Wirth, Aron B. Bender, David M. German, Ryle Przybylowicz, Peter van Dam, Thomas A. Dewland, Khidir Dalouk, Eric Stecker, Babak Nazer, Peter M. Jessel, Karen S. MacMurdy, Ignatius Gerardo E. Zarraga, Bassel Beitinjaneh, Charles A. Henrikson, Merritt Raitt, Cristina Fuss, Maros Ferencik, Larisa G. Tereshchenko

## Abstract

**Introduction:** Adaptive cardiac resynchronization therapy (aCRT) is known to have clinical benefits over conventional CRT; however, their effects on the electrical dyssynchrony have not been compared.

**Methods:** We conducted a double-blind, randomized controlled trial in patients receiving CRT for routine clinical indications. Participants underwent cardiac computed tomography and 128-electrodes body surface mapping. We measured electrical dyssynchrony on the epicardial surface using noninvasive electrocardiographic imaging (ECGI) before and 6 months post-CRT. Ventricular electrical uncoupling (VEU) was calculated as the difference between the mean left ventricular (LV) and right ventricular (RV) activation times. An electrical dyssynchrony index (EDI) was computed as the standard deviation of local epicardial activation times.

**Results:** We randomized 27 participants (mean age 64±12 y; 34% female; 53% ischemic cardiomyopathy; LV ejection fraction 28±8%; QRS duration 155±21 ms; strict left bundle branch block (LBBB) in 13%). In atypical LBBB (n=11;41%) with S-waves in V_5_-V_6_, conduction block occurred in the anterior RV, as opposed to the interventricular groove in those who met the strict LBBB criteria. As compared to baseline, VEU reduced post-CRT in aCRT (median reduction 18.9(interquartile range 4.3-29.2 ms; P=0.034), but not in conventional CRT (21.4(−30.0 to 49.9 ms; P=0.525) group. There were no differences in the degree of change in VEU and EDI indices between treatment groups.

**Conclusion:** The effect of aCRT and conventional CRT on electrical dyssynchrony is largely similar. Further studies are needed to investigate if atypical LBBB with prominent S wave in V_5_-V_6_ responds to His bundle pacing.

## Introduction

The adaptive cardiac resynchronization therapy (aCRT) algorithm has been developed to optimize CRT delivery by avoiding right ventricular (RV) pacing and providing concomitant dynamic atrioventricular (AV) and ventriculo-ventricular (VV) adjustments through the measurement of intracardiac conduction parameters.^1^ A randomized controlled trial comparing aCRT and conventional CRT showed that aCRT is safe and at least as effective as conventional CRT.^2^ A higher percentage of left ventricular (LV) pacing was associated with superior clinical outcomes, including decreased risk of death, heart failure (HF) hospitalizations,^3^ and 30-day readmissions.^4^ Further analysis comparing aCRT to the historic conventional CRT controls showed that the proportion of clinical responders to aCRT was higher by 12%.^5^ Notably, aCRT reduced the occurrence of atrial fibrillation (AF) by continuously adjusting AV intervals.^6-9^

However, aCRT’s impact on electrical dyssynchrony is not entirely clear. Experimental studies in left bundle branch block (LBBB)-failing hearts showed that bi-ventricular (BiV) pacing, but not LV pacing, decreased electrical dispersion.^10^ Some previous randomized controlled trials that compared LV and BiV pacing showed a similar benefit for BiV and LV CRT.^11-13^ Further measurements aCRT’s effect on electrical dyssynchrony, such as epicardial activation map analysis and novel vectorcardiographic (VCG) metrics, have not previously been assessed. A better understanding of the mechanisms behind aCRT is essential for further improvement in CRT delivery and clinical outcomes.

To investigate the effect of aCRT on electrical dyssynchrony, we conducted a randomized controlled trial (RCT) “Adaptive CRT Effect on Electrical Dyssynchrony” (aCRT-ELSYNC; ClinicalTrials.gov Identifier: NCT02543281). We hypothesized that (1) aCRT (as compared to conventional CRT) provides a superior degree of reduction of electrical dyssynchrony 6-months post-CRT, and (2) surface ECG/VCG metric, sum absolute QRST integral (SAIQRST),^14, 15^ is a superior estimate of electrical dyssynchrony as compared to QRS duration. The secondary objective was to determine if aCRT is associated with improvement in the quality of life and other clinically important outcomes.

## Method

### Trial design

We conducted a single-center, double-blind, 1:1 parallel-group assignment RCT. The study was approved by the Oregon Health & Science University (OHSU) Institutional Review Board (IRB), and it was monitored by the Data and Safety Monitoring Board. All study participants signed written informed consent before entering the study. Study data were collected and managed using REDCap electronic data capture tools hosted at OHSU.^16, 17^

Six weeks after trial commencement, the registry arm was added to include the study participants who failed the RCT eligibility criteria screening. Furthermore, in the fall of 2018, an initial exclusion criterion (previous or existing CRT system, implantable cardioverter-defibrillator (ICD), or pacemaker) was broadened to include individuals with a previously implanted single-chamber ICD if they had a documented history of ventricular pacing (VP) <1% and an indication for CRT upgrade.

There were two study visits. The first was prior to CRT device implantation. The second visit was six months post-CRT implantation. Both visits included documentation of the medical history and use of medications, the quality of life assessment using the 36-Item Short Form Health Survey (SF-36) and Minnesota Living with Heart Failure Questionnaire (MLHFQ), recording of 12-lead Holter ECG at rest and during a 6-minute walk test, and body surface potential mapping. Cardiac computed tomography (CT) scan or cardiac magnetic resonance (CMR) study was performed once, before the device implantation. In addition, a copy of digital intracardiac electrograms was saved during CRT implantation.

Echocardiography data and assessment of the New York Heart Association (NYHA) class data were obtained from the medical records.

#### Eligibility criteria for study participants

All OHSU patients who were scheduled for CRT device implantation at OHSU were considered for participation.

Inclusion criteria were standard class I or class II indications for CRT pacemaker (CRT-P) or CRT-defibrillator (CRT-D) implantation in accordance with ACC/AHA/HRS guidelines,^18^ age ≥ 18 years, and willingness and ability to comply with the protocol.

Exclusion criteria were:

1. Presence of a permanent AF. AF was considered permanent when it was continuously present, and the physician and patient decided not to make any further attempts at rhythm control.
2. Previous/existing CRT system or pacemaker, or existing ICD if the amount of ventricular pacing was >1%.
3. Estimated glomerular filtration rate (eGFR) < 30 ml/min.
4. Unstable angina, acute myocardial infarction (MI), coronary artery bypass graft (CABG) surgery, or percutaneous coronary intervention (PCI) within 30 days before enrollment.
5. Valvular heart disease with indications for valve repair or replacement.
6. Enrollment in any other interventional trial.
7. Pregnancy.
8. Post-heart transplant status.
9. New NYHA class IV HF or progression to NYHA class IV HF within three months before enrollment.
10. Concomitant condition other than a cardiac disease that is associated with a higher likelihood of death during one year after enrollment.
11. Inability or unwillingness to cooperate or give written informed consent.

#### Intervention

The study participants were implanted with a commercially available, clinically indicated Medtronic CRT device with aCRT algorithm (Viva™ models DTBA1D4, DTBA1D1, DTBA1Q1, DTBA1QQ, DTBB1Q1, DTBB1QQ, DTBB1D1, DTBB1D4, or C6TR01). Any compatible lead could be implanted. LV lead location was selected at the discretion of the implanting electrophysiologist. The aCRT intervention arm had adaptive BiV and LV pacing programmed ON, while the conventional CRT control arm had nonadaptive CRT programmed ON.

#### Randomization and blinding

Study participants were randomized using a random stratified block, with assignment ratio 1:1, and two assignment strata: by sex (male vs. female) and cardiomyopathy type (ischemic vs. nonischemic). We used a random block size (2, 4, or 6) with allocation proportional to the elements of Pascal’s triangle. A random seed was used, and the randomization list was created using the *–ralloc-* program in Stata version 13.1.

Randomization was enacted via sealed envelopes containing randomization assignments delivered to a Medtronic field representative/device technician who programmed the CRT device immediately after the implantation. The participants, implanting team, and study investigators assessing outcomes were blinded to the intervention.

#### Primary endpoints

For the 1^st^ hypothesis, the primary endpoint was defined as a change in electrical dyssynchrony metrics measured on epicardial activation map and surface ECG six months post-CRT, as compared to pre-CRT.

For the 2nd hypothesis, the primary endpoint was defined as a correlation between the surface ECG metrics of dyssynchrony (SAIQRST, in comparison to QRS duration and QRS area) and electrical dyssynchrony metrics measured on the epicardial activation map, assessed prior to CRT implantation.

### Building the epicardial activation map

We employed an electrocardiographic imaging (ECGI)^19, 20^ approach to build an epicardial activation map.^21^ For each study participant, we built two maps: before CRT implantation and six months post-CRT. Figure 1 shows the steps in data analysis. To obtain the solution of the inverse problem, we used the SCIRun problem-solving software developed at the Center for Integrative Biomedical Computing (University of Utah, UT, US)^22^.

**Figure 1:**
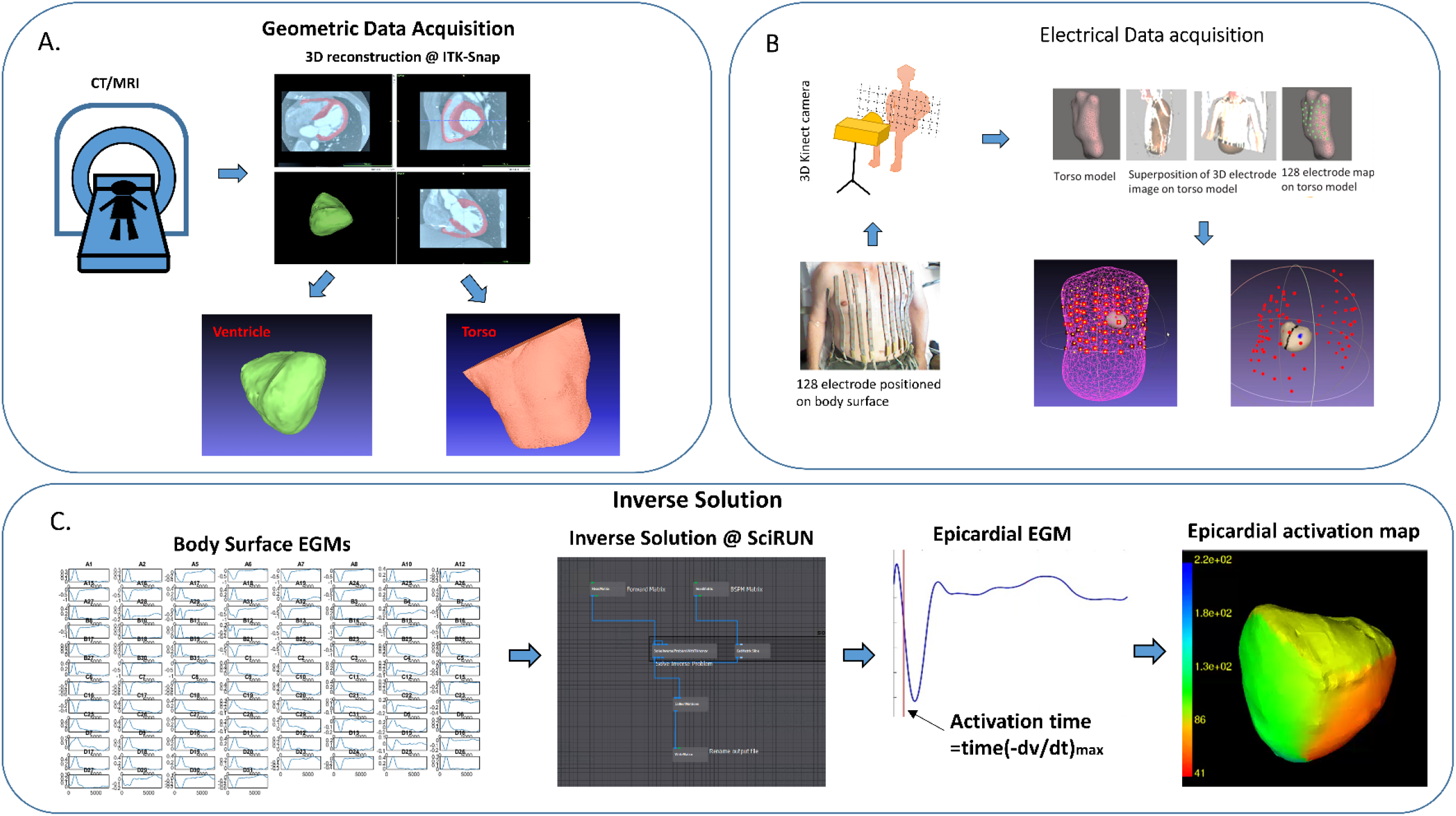
Study workflow. (**A**) CT scan or CMR provided the geometry of the ventricles and the torso. Images underwent segmentation, structure identification, geometric modeling and meshing. The 3D meshes of the ventricles and torso were created. (**B**) Body surface potential (BSP) was recorded using the 128 electrodes. 3D photography was used to record the electrodes locations on a torso. The torso geometric models based on the 3D photography and CT/CMR were co-registered. (**C**) Epicardial EGM reconstruction using the SCIRun inverse solution module. Local activation for each epicardial node was obtained as the point of the steepest downward slope (minimum dV/dt) of the corresponding EGM. An epicardial activation map was generated.

The study participants underwent recording of unipolar ECG potentials on the body surface using the ActiveTwo biopotential measurement system (BioSemi, Amsterdam, the Netherlands) with 128 Ag/AgCL electrodes (4 panels of 32 electrodes; each panel is arranged as four strips of 8-electrodes; diameter of the ECG electrodes 5 mm),^23^sampling rate 16,384 Hz and bandwidth DC-3,200 Hz. The localization of the electrodes was performed using a three dimensional (3D) photography approach using PeacsKinect (Peacs BV, Arnhem, the Netherlands), as previously described.^23^ Immediately after the body potentials recording, participants underwent cardiac imaging while wearing ≥ 5 electrode markers for subsequent co-registration of torso images. Body surface potentials were recorded twice: before CRT implant and 6 months post-implant.

Before CRT implantation, study participants underwent either non-contrast CT or CMR study. Non-contrast CMR used a Siemens TIM Trio 3 Tesla with VB17 software and Siemens Prisma Fit 3 Tesla scanner with E11C software. Prospectively ECG-triggered non-contrast 128-slice cardiac CT (Philips iCT SP, Philips Medical Imaging, Cleveland, OH) images were acquired in mid to end-diastole with a slice thickness of 0.6 mm and in-plane resolution of ~0.5 mm. Two cardiologists (RP, MF) reviewed cardiac CT/CMR images and identified interventricular and atrioventricular grooves. Ventricular volumes were obtained in a semiautomatic fashion using commercially available software (IntelliSpace Portal; Philips Healthcare, Redmond, WA, US for CT; and CVI42; Circle Cardiovascular Imaging Inc., Calgary, Alberta, Canada for MRI).

We used the collected CT/MRI data to reconstruct the 3D mesh of the ventricle and torso. A semi-automated mesh generation algorithm^23^ (ITK-snap software, PICSL, United States)^24^ was applied to obtain the meshes (Figure 1). Each cardiac mesh was manually reviewed (KTH) to ensure a continuous segmentation of the epicardium of both ventricular chambers and exclude atria chambers. The torso meshes were also manually reviewed (KTH).

A normal sinus beat (baseline) or a paced beat (follow up) was selected for analyses. Wilson’s central terminal (WCT) potential was calculated by averaging the potentials recorded by the electrodes placed in the three extremities (left arm, right arm, and left leg). The unipolar voltage at each of 128 torso electrodes was then calculated by subtracting WCT potential from the potential recorded by the torso electrode.

We used the boundary element solution provided by the ‘BuildBEmatrix’ in the forwardinverse toolkit in SCIRun. BuildFEMatrix inputs a finite element mesh with the conductivities set on each element. The output transfer matrix is a stiffness matrix based on the Galerkin method. The module used the ventricular mesh as the ‘inner surface’ and the surface built by *n* number of electrodes on the torso as the ‘outer surface.’ Once the transfer matrix A was created, the next step was to solve the inverse problem for the epicardial potentials using the zero-order Tikhonov regularization scheme provided by the ‘Solve lnverse Problem With Tik honov’ in the SCIrun forward-inverse toolkit. We used the manual selection option provided in the module to select a range of solution constraint parameter γ values. The selected γ values warranted the formation of the “L” shaped curve (known as “L-curve” approach) when the residual norm and the weighted solution norm was plotted in a log-log plot. The resulting solution matrix provided the calculated unipolar epicardial electrograms (EGMs) at each node of the epicardial mesh. The resolution of the ventricular mesh was 3.2±1.1 mm, with 3,961±658 nodes.

Local activation time (LAT) was calculated as the time between a common reference point (body surface potential QRS onset) and the point of steepest downward slope (minimum dV/dt) of the epicardial EGM on each epicardial node within one-fifth of the cardiac cycle measured from the local absolute maximum of unipolar EGM signal (Figure 1C). Accuracy of automated LAT detection was manually verified by three investigators (KTH, KJL, AW) on a random sample of EGMs for each map. If inaccurate LAT was due to erroneous minimum dV/dt detection, it was corrected. If an epicardial EGM did not display a morphology allowing a single accurate LAT identification, the corresponding epicardial node was removed.

### Electrical Dyssynchrony on Epicardial Activation Maps

Electrical dyssynchrony on the epicardial activation map was quantified by the following metrics.^25, 26^ The RV activation time (RVAT) was defined as the time between the earliest and the latest site of RV activation. The LV activation time (LVAT) was defined as the time between the earliest and the latest site of LV activation. The global total activation time (TAT) was defined as the time between the earliest and the latest site of the total ventricular epicardial activation. Ventricular electrical uncoupling (VEU) was measured as the difference between the mean LV and RV activation times. Positive VEU indicated LV uncoupling (delay) from the RV, whereas negative VEU indicated RV uncoupling (delay) from the LV.^26^ An electrical dyssynchrony index (EDI) was computed as the standard deviation of activation times throughout the entire ventricular epicardium (EDI_V_), LV epicardium (EDI_LV_), and RV epicardium (EDI_RV_).

### Electrical Dyssynchrony Indices on Surface ECG and VCG

Assessment of the baseline QRS duration and morphology (ventricular conduction abnormality) was performed using clinical 12-lead ECG recorded before CRT implant, stored in the electronic medical record.

For vectorcardiographic (VCG) analysis, to ensure adequate comparison of electrical dyssynchrony measured on surface ECG and epicardial activation maps, we constructed a 12-lead ECG using recorded body surface potentials and analyzed the same cardiac beat. We identified the location of the V_1_-V_6_ electrodes in the nearest proximity of the expected precordial leads location (Supplemental Figure 1). Limb lead I was calculated as the difference of left arm – right arm potentials. Lead II was calculated as the difference of left leg – right arm potentials, and lead III is the difference of left leg – left arm potentials. Kors transformation matrix was used to transform the 12-lead to orthogonal XYZ ECG. Per study protocol, electrical dyssynchrony on surface ECG was measured by QRS duration and sum absolute QRST integral (SAIQRST).^14, 15^ In addition, we measured QRS duration and QRS area^27^ as previously described.^28, 29^

### Secondary clinical endpoints

The secondary clinical endpoints were assessed by the research team members blinded to the randomization assignment. The clinical composite outcome was defined as worsened if a study participant (1) died from any cause, or (2) experienced HF hospitalization within six months post-CRT. In addition, secondary endpoints included quality of life assessment by the Minnesota Living with Heart Failure Questionnaire (MLHFQ) and 36-Item Short Form Survey (SF-36) questionnaire. The 6-minute walk distance was measured.

### Statistical analysis

The planned sample size (32 participants) was estimated based on prior observational study data,^30^ reporting the distribution of EDI in CRT responders and non-responders.

We conducted an intention-to-treat analysis. Wilcoxon (Mann-Whitney) rank-sum exact test was used to compare continuous variables that were summarized as the median and interquartile range (IQR). Fisher’s exact test was used to compare categorical variables. Wilcoxon matched-pairs signed-rank test was used to compare changes from the baseline to 6-months post-CRT. Spearman’s correlation statistics were computed to study an association between epicardial and surface ECG measures of dyssynchrony. Two-sided exact P-values were reported. *P*<0.05 was considered statistically significant. Statistical analyses were performed using STATA MP 16.1 (StataCorp LLC, College Station, TX).

## Results

### Study Population

The study flowchart is shown in Figure 2. Enrollment in the study was completed between June 22^nd^, 2015, and October 10^th^, 2018. All study participants had LV lead successfully implanted within the epicardial venous system. Out of 32 enrolled participants, 27 were randomized, and five were included in the registry arm. All randomized participants, and 4 out of the 5 registry participants completed the 6-month follow-up visit procedures.

**Figure 2.**
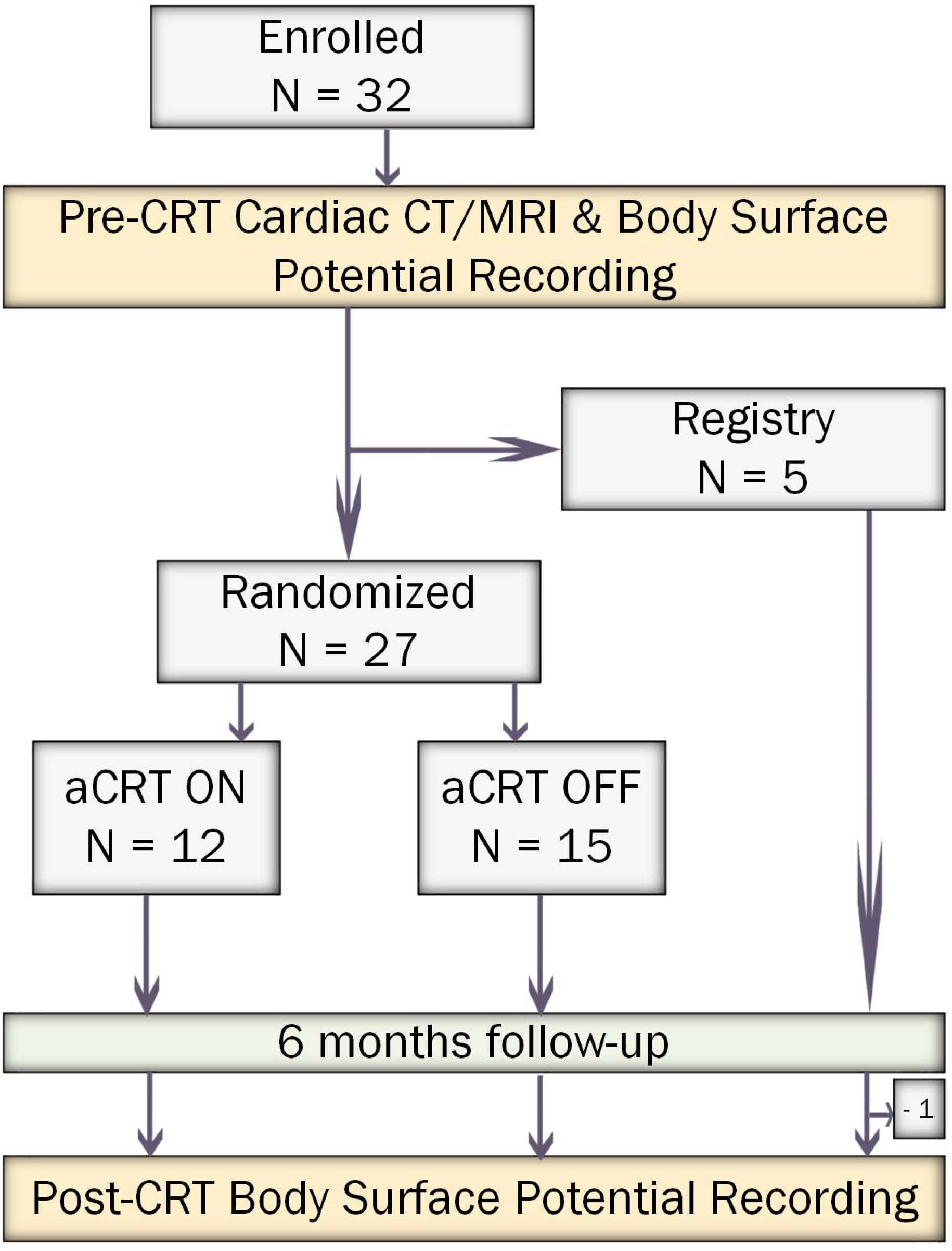
Study flowchart.

The clinical characteristics of the study participants are shown in Table 1. One-third of the participants were female, and approximately half had ischemic cardiomyopathy. Out of randomized participants, 15% (n=4) met strict LBBB criteria (broad notched or slurred R without Q and S waves in leads I, aVL, V_5_, and V_6_; intrinsicoid deflection in V_5_-V_6_ > 60ms), 41% (n=11) met LBBB criteria used in the main CRT trials (RsR’ in V_6_ and rS/QS in V_1_-V_2_) and had prominent S waves in V_5_-V_6_, and 44% (n=12) had IVCD with QRS duration above 150 ms. Nearly all participants were on ACEI or ARBs and beta-blockers. Study groups were well-balanced.

**Table 1:** Baseline clinical and demographic characteristics of the study participants.

| **Characteristic** | **All (n=32)** | **Conventional CRT (n=15)** | **Adaptive CRT (n=12)** | **Registry (n=5)** |
| --- | --- | --- | --- | --- |
| Age(SD), y | 64.4(11.5) | 65.3(12.2) | 62.2(10.3) | 67.3(13.6) |
| Female, n(%) | 11(34.4) | 5(33.3) | 4(33.3) | 2(40.0) |
| White, n(%) | 31(96.9) | 15(100) | 11(91.7) | 5(100.0) |
| Body mass index(SD), kg/m<sup>2</sup> | 31.3(7.4) | 30.2(4.3) | 33.1(9.1) | 30.2(10.8) |
| Ischemic cardiomyopathy, n(%) | 17(53.1) | 9(60.0) | 5(41.7) | 3(60.0) |
| Myocardial infarction history, n(%) | 9(28.1) | 4(26.7) | 3(25.0) | 2(40.0) |
| Revascularization history, n(%) | 15(46.9) | 8(53.3) | 5(41.7) | 2(40.0) |
| 3-vessel disease, n(%) | 8(25.0) | 4(26.7) | 3(25.0) | 1(20.0) |
| NYHA class II, n(%) | 21(65.6) | 9(60.0) | 8(66.7) | 4(80.0) |
| NYHA class III, n(%) | 11(34.4) | 6(40.0) | 4(33.3) | 1(20.0) |
| QRS duration(SD), ms | 155.1(20.9) | 151.3(24.3) | 160.6(19.0) | 153.1(20.9) |
| QTc interval(SD), ms | 492.5(40.6) | 492.0(45.0) | 496.1(35.1) | 485.6(47.2) |
| PR interval(SD), ms | 181.2(22.9) | 187.6(21.6) | 176.6(21.0) | 170.0(35.4) |
| Strict LBBB, n(%) | 4(12.5) | 1(6.7) | 3(25.0) | 0 |
| IVCD, n(%) | 28(87.5) | 14(93.3) | 1(75.0) | 5(100.0) |
| Upgrade from ICD, n(%) | 1(3.1) | 0 | 1(8.3) | 0 |
| Atrial fibrillation history, n(%) | 7(21.9) | 3(20.0) | 2(16.7) | 2(40.0) |
| Diabetes, n(%) | 7(21.9) | 4(26.7) | 2(16.7) | 2(20.0) |
| Hypertension, n(%) | 24(75.0) | 11(73.3) | 8(66.7) | 5(100.0) |
| COPD, n(%) | 4(12.5) | 3(20.0) | 1(8.3) | 0 |
| Never-smoker, n(%) | 20(62.5) | 7(46.7) | 8(66.7) | 5(100.0) |
| Beta-blockers, n(%) | 31(96.9) | 14(93.3) | 12(100.0) | 5(100.0) |
| ACEI or ARB, n(%) | 28(87.5) | 13(86.7) | 10(83.3) | 5(100.0) |
| LVEF(SD), % | 28.4(8.0) | 26.0(8.6) | 29.7(7.5) | 32.3(6.4) |
| LVDDi(SD), cm/m<sup>2</sup> | 3.2(0.5) | 3.3(0.6) | 3.1(0.4) | 3.1(0.3) |
| LVSDi(SD), cm/m<sup>2</sup> | 2.7(0.5) | 3.0(0.5) | 2.7(0.4) | 2.6(0.2) |

SD=standard deviation; NYHA=New York Heart Association; ICD=implantable cardioverter-defibrillator; LBBB=left bundle branch block; IVCD=interventricular conduction delay; COPD=chronic obstructive pulmonary disease; ACEI=angiotensin-converting enzyme inhibitor; ARB=angiotensin receptor blocker; LVEF=left ventricular ejection fraction; LVDDi=left ventricular diastolic dimension indexed to body surface area; LVSDi=left ventricular systolic dimension indexed to body surface area.

### Effect of aCRT on ventricular epicardial activation maps and electrical dyssynchrony

At baseline, epicardial activation in a strict LBBB (Figure 3A and Supplemental Figure 2) manifested with RV breakthrough that began in the mid-RV and rapidly activated the entire RV, whereas LV activation was delayed. The line of conduction block corresponded to the interventricular groove. Epicardial activation in atypical LBBB with S-waves in V_5_-V_6_ (Figure 4A and Supplemental Figure 3) differed from typical LBBB in two ways. First, the line of the conduction block was located more anteriorly, away from the interventricular groove, in the RV vicinity. Second, the epicardial activation spreading from RV to LV involved apical LV segments before or at the same time as basal LV segments. Overall, the LV activation patterns were remarkably variable in study participants.

**Figure 3:**
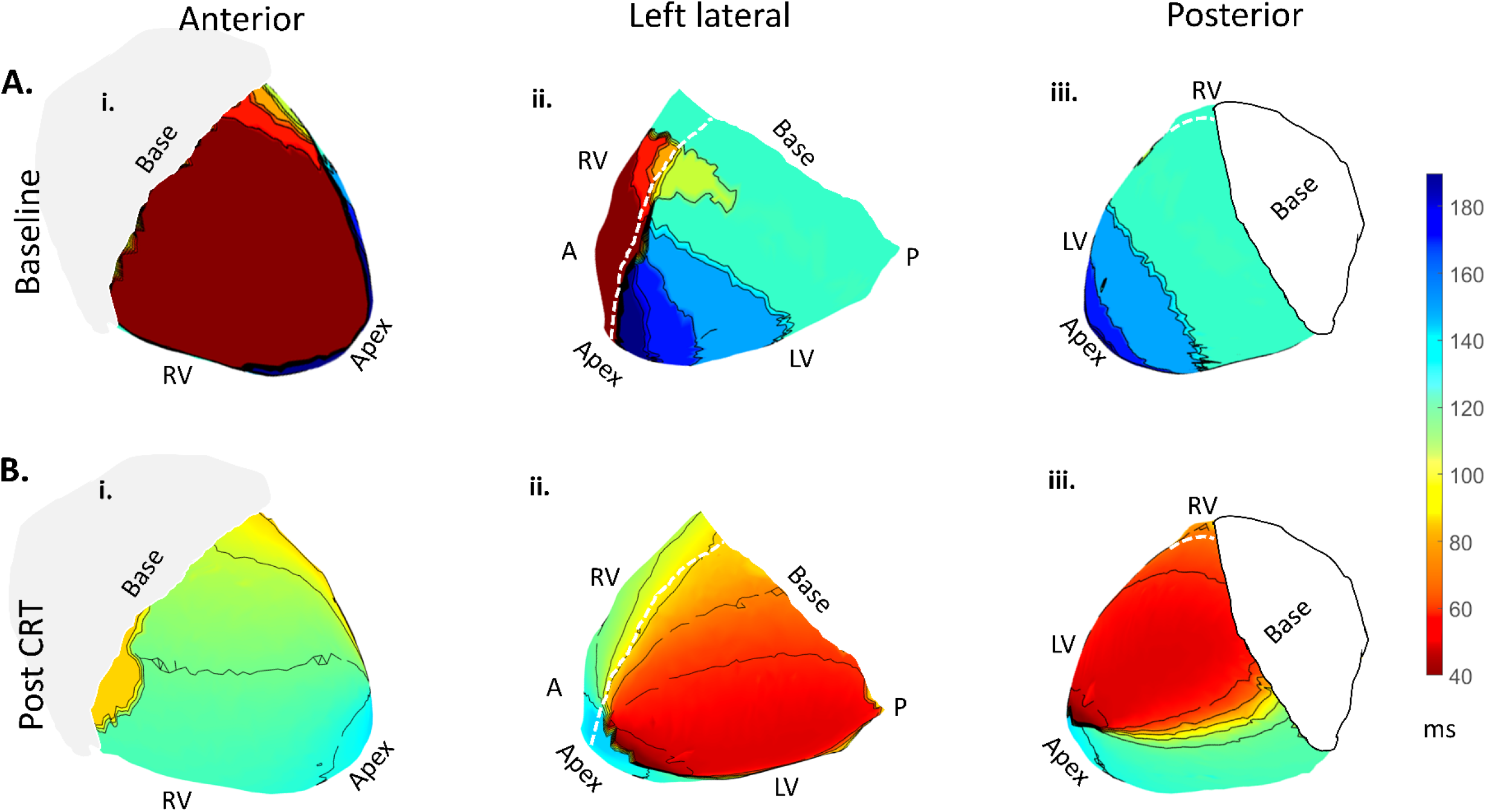
Representative example of epicardial activation in a female participant with NICM and typical LBBB at baseline (**A**) and 6-month post-aCRT in the treatment arm (**B**) in (**i**) anterior, (**ii**) left lateral, and (**iii**) posterior views. A white dashed line marks the interventricular groove. Epicardial 10-ms isochrones show local activation time (LAT), measured from the surface ECG QRS onset to minimum dV/dt, color-coded from red (early) to blue (late). Thick black lines (crowded isochrones) indicate line/region of conduction block.

LV-only pacing in the aCRT arm presented with the earliest activation in the LV epicardium and gradual propagation of activation (Figure 3B). Bi-V pacing in the conventional CRT arm (Figure 4B) had two early activation centers, corresponding to RV and LV leads. Most lines of conduction block previously observed in sinus rhythm shifted or disappeared during ventricular pacing, suggesting the functional nature of the block (Figures 3-4). The electrical activation sequence was patent-specific.

**Figure 4:**
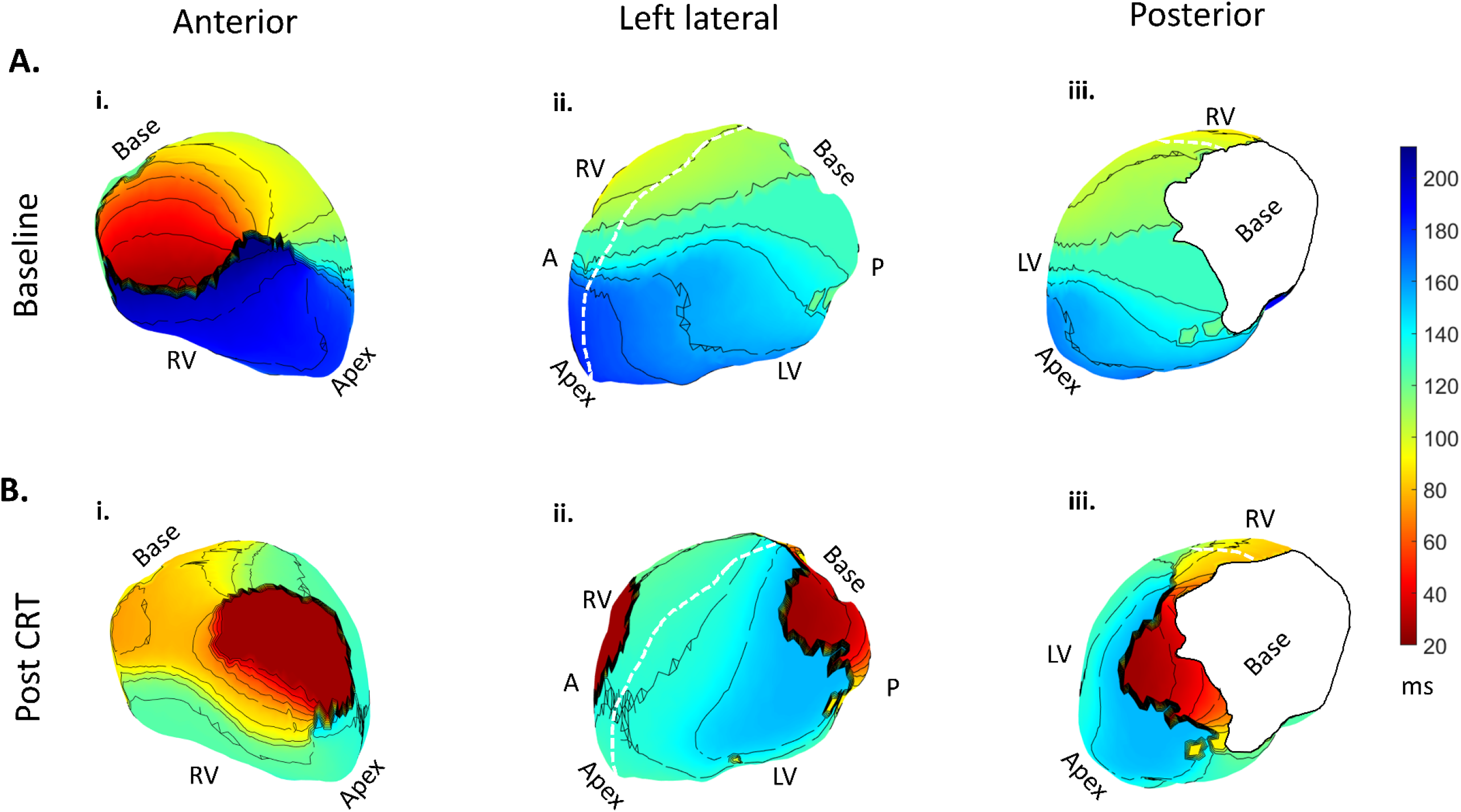
Representative example of epicardial activation in a female participant with NICM and IVCD (atypical LBBB with S in V_5_-V_6_) at baseline (**A**) and 6-month post-BiV CRT in the control arm (**B)** in (**i**) anterior, (**ii**) left lateral, and (**iii**) posterior views. See Figure 3 legend for details.

At baseline, there were no differences in the dyssynchrony metrics between treatment groups (Table 2). Post-CRT, TAT, LVAT, and RVAT were reduced significantly and similarly in both groups. Baseline VEU ranged from negative 85 to positive 72 ms, indicating that the study participants had dyssynchrony due to both LV and RV uncoupling, without significant differences between treatment groups. Post-CRT, VEU significantly decreased only in the aCRT group, but not in the conventional CRT arm. Post-CRT, aCRT significantly reduced RV uncoupling (Table 2). This is illustrated by changes in VEU in each study participant (Figure 5). Harmonious VEU reduction was observed in aCRT group, in contrast to the conventional CRT arm, demonstrating discordant and inconsistent changes in VEU. However, there was no statistically significant difference in the degree of VEU reduction between treatment groups.

**Table 2:** Comparison of ventricular epicardial activation metrics of electrical dyssynchrony.

| | Conventional CRT (n=15) | $P_{pre-post}$ | Adaptive CRT (n=12) | $P_{pre-post}$ | $P$-value |
| --- | --- | --- | --- | --- | --- |
| Baseline TAT median(IQR),ms | 192(154-213) | - | 190(172-215) | - | 0.801 |
| Post-CRT TAT median(IQR),ms | 111(94-126) | - | 101(98-134) | - | 0.674 |
| Difference TAT median(IQR),ms | -69(-54 to -124) | 0.0002 | -78(-54 to -111) | 0.0005 | 0.857 |
| Baseline LVAT median(IQR),ms | 172(138-213) | - | 187(172-213) | - | 0.622 |
| Baseline mean RV time median(IQR), ms | 169(127-198) |  | 158(142-184) |  | 0.829 |
| Post-CRT mean RV time median(IQR), ms | 105(91-133) |  | 107(100-148) |  | 0.347 |
| Difference mean RV time median(IQR), ms | -48(0107 to -3) | 0.003 | -47(-59 to -23) | 0.002 | 0.614 |
| Baseline mean LV time median(IQR), ms | 160(136-214) |  | 132(121-151) |  | 0.083 |
| Post-CRT mean LV time median(IQR), ms | 106(79-141) |  | 108(93-132) |  | 0.905 |
| Difference mean LV time median(IQR), ms | -50(-83 to -14) | 0.003 | -27(-36 to -8) | 0.043 | 0.183 |
| Baseline LVAT median(IQR),ms | 172(138-213) | - | 187(172-213) | - | 0.622 |
| Post-CRT LVAT median(IQR),ms | 104(87-126) | - | 100(93-131) | - | 0.895 |
| Difference LVAT median(IQR),ms | -58(-36 to -124) | 0.0004 | -75(-48 to -110) | 0.0005 | 0.782 |
| Baseline RVAT median(IQR),ms | 187(141-211) | - | 185(153-202) | - | 0.895 |
| Post-CRT RVAT median(IQR),ms | 110(87-126) | - | 102(89-125) | - | 0.783 |
| Difference RVAT median(IQR),ms | -60(-22 to -111) | 0.0006 | -83(-60 to -91) | 0.0005 | 0.406 |
| Baseline VEU median(IQR),ms | +2.3(-34.8 to 17.2) |  | -28.4(-40.3 to -11.2) |  | 0.183 |
| Post-CRT VEU median(IQR),ms | -3.3(-13.3 to 18.0) |  | -9.1(-18.2 to 4.7) |  | 0.347 |
| Difference VEU median(IQR),ms | 21.4(-30.0 to 49.9) | 0.525 | 18.9(4.3 to 29.2) | 0.034 | 0.719 |
| Baseline EDI<sub>V</sub> median(IQR), ms | 52.5(41.8-68.5) | - | 53.1(40.2-69.2) | - | 0.792 |
| Post-CRT EDI<sub>V</sub> median(IQR), ms | 30.6(23.6-45.2) | - | 30.8(22.1-46.1) | - | 0.683 |
| Difference EDI<sub>V</sub> median(IQR), ms | -12.9(-6.6 to -38.9) | 0.0001 | -16.4(-8.1 to -40.1) | 0.0005 | 0.755 |
| Baseline EDI<sub>LV</sub> median(IQR), ms | 50.1(32.8-73.2) | - | 47.5(40.6-64.2) | - | 0.981 |
| Post-CRT EDI<sub>LV</sub> median(IQR), ms | 31.6(23.5-42.0) | - | 31.1(25.1-44.0) | - | 0.943 |
| Difference EDI<sub>LV</sub> median(IQR), ms | -11.6(-0.2 to -51.2) | 0.0002 | -13.1(05.6 to -35.1) | 0.0005 | 0.943 |
| Baseline EDI<sub>RV</sub> median(IQR),ms | 37.0(35.5-67.7) | - | 44.4(28.8-59.4) | - | 0.867 |
| Post-CRT EDI<sub>RV</sub> median(IQR),ms | 29.2(21.9-41.1) | - | 22.6(17.3-39.0) | - | 0.347 |
| Difference EDI<sub>RV</sub> median(IQR),ms | -17.7(+8.3 to -37.8) | 0.0001 | -20.3(-9.3 to -32.7) | 0.0005 | 0.943 |

$P_{pre-post}$ = $P$ -value for difference baseline – post-CRT

**Figure 5.**
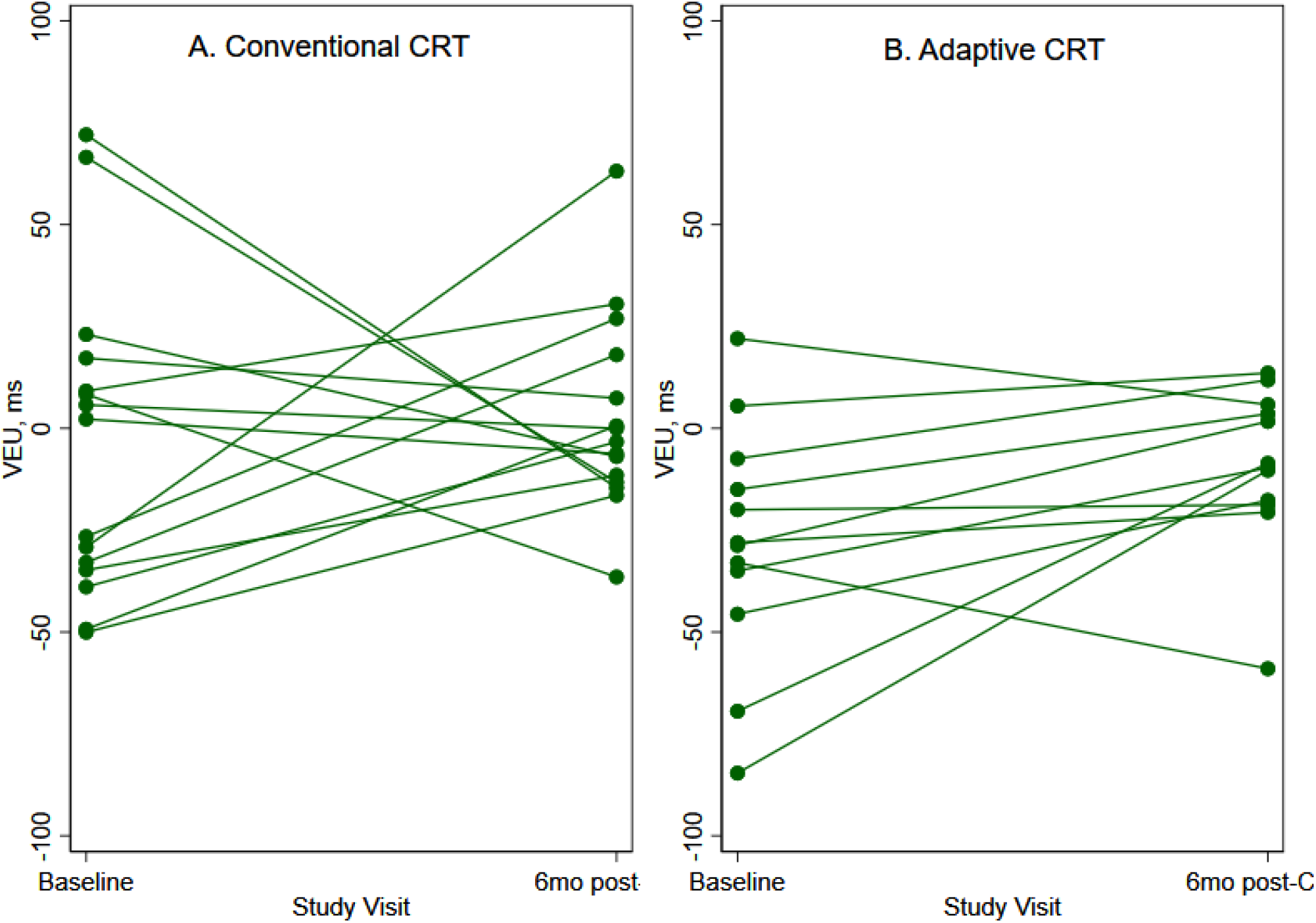
Change in the ventricular electrical uncoupling (VEU) between two study visits (green line) in every randomized study participant (green dot), in (A) conventional CRT, and (B) adaptive CRT arms.

Both aCRT and conventional CRT resulted in a significant reduction of both intra-ventricular and inter-ventricular dyssynchrony as measured by EDI indices, without differences between treatment groups (Table 2).

### Effect of aCRT on surface ECG and VCG

QRS area and SAIQRST significantly decreased post-CRT in both groups, whereas QRS duration changes were borderline. However, no significant differences were found in QRS duration, QRS area, and SAIQRST between the two treatment groups at baseline or after therapy (Table 3).

**Table 3:** Comparison of surface ECG metrics of electrical dyssynchrony.

| | Conventional CRT (n=15) | $P_{pre-post}$ | Adaptive CRT (n=12) | $P_{pre-post}$ | $P$-value |
| --- | --- | --- | --- | --- | --- |
| Baseline QRS duration median(IQR), ms | 156(135-174) | - | 166(145-172) | - | 0.379 |
| Post-CRT QRS duration median(IQR), ms | 139(124-158) | - | 136(120-149) | - | 0.783 |
| Difference QRS duration median(IQR), ms | -9(-31 to 2) | 0.076 | -31.5(-38 to -19) | 0.043 | 0.128 |
| Baseline SAIQRST median(IQR), mVms | 210(142-299) | - | 225(154-405) | - | 0.456 |
| Post-CRT SAIQRST median(IQR), mVms | 126(106-189) | - | 143(103-182) | - | 0.943 |
| Difference SAIQRST median(IQR), mVms | -49(-108 to -19) | 0.005 | -105(-254 to -22) | 0.012 | 0.427 |
| Baseline VM QRS area median(IQR), mVms | 243(202-414) | - | 353(215-647) | - | 0.548 |
| Post-CRT VM QRS area median(IQR), mVms | 190(152-257) | - | 156(137-263) | - | 0.546 |
| Difference VM QRS area median(IQR), mVms | -82(-250 to 10) | 0.030 | -215(-458 to 17) | 0.021 | 0.323 |

$P_{pre-post}$ = $P$ -value for difference baseline – post-CRT; VM=vector magnitude.

### Correlation of the dyssynchrony measured on surface ECG and epicardial surface

There was no statistically significant correlation between any of the epicardial and surface ECG measures of dyssynchrony was found in all (including registry) study participants at baseline (Supplemental Table 1).

### Secondary outcomes

There were no deaths and HF hospitalizations during 6 months of follow-up. There were no differences in the 6-minute walk distance between treatment groups. Baseline and post-CRT 6-minute walk distance did not differ either (Table 4). Overall, MLHFQ score improved from baseline [median 46(IQR 20-68)] to post-CRT [median 27(IQR 11-48); P=0.003]. However, there were no differences between treatment groups.

**Table 4:** Quality of life and 6-minute walk distance changes.

|  | Conventional CRT (n=15) | Adaptive CRT (n=12) | P-value |
| --- | --- | --- | --- |
| Baseline 6-min walk distance median(IQR), m | 360(312-374) | 362(345-438) | 0.358 |
| Post-CRT 6-min walk distance median(IQR), m | 352(318-412) | 368(328-402) | 0.820 |
| Difference 6-min walk distance median(IQR), m | +18(-12 to +41) | -19(-76 to +57) | 0.804 |
| Baseline MLHFQ total score median(IQR) | 56(20-70) | 33(15-65) | 0.373 |
| Post-CRT MLHFQ total score median(IQR) | 30(11-50) | 21(8-44) | 0.478 |
| Difference MLHFQ total score median(IQR) | -20(-30 to -4) | -7(-15 to 0.5) | 0.353 |
| Difference SF-36 physical functioning median(IQR) | 0(0 to +25) | +10(-5 to +18) | 0.970 |
| Difference SF-36 role physical median(IQR) | 0(0 to +25) | +13(0 to +38) | 0.409 |
| Difference SF-36 role emotional median(IQR) | 0(0 to 0) | 0(0 to +50) | 0.724 |
| Difference SF-36 energy/fatigue median(IQR) | +10(+5 to +35) | +8(-10 to +18) | 0.255 |
| Difference SF-36 emotional median(IQR) | +4(-8 to +16) | 0(-6 to +16) | 0.929 |
| Difference SF-36 social functioning median(IQR) | +13(0 to +25) | 0(0 to +6) | 0.396 |
| Difference SF-36 pain score median(IQR) | +13(-10 to +23) | 0(-6 to +11) | 0.364 |
| Difference SF-36 general health median(IQR) | +10(-5 to +10) | -5(-8 to 10) | 0.474 |

On average, role limitations due to physical health improved with CRT (from median 0 (IQR 0-63) to median 100 (IQR 0-100); P=0.004). The energy/fatigue score improved from a median 30(IQR 20-53) to median 55(IQR 35-70); P=0.004. There was no pre-versus post-CRT difference in other SF-36 indicators and no difference in any SF-36 indicators between treatment groups (Table 4).

## Discussion

Our double-blind RCT showed that the effect of aCRT and conventional CRT on electrical dyssynchrony is largely similar, with both significantly reducing electrical dyssynchrony overall. We did not find differences between aCRT and conventional CRT in the degree of reduction of LV and RV inter- and intraventricular electrical dyssynchrony 6-month post-CRT. However, only aCRT but not conventional CRT harmoniously reduced interventricular dyssynchrony by reducing RV uncoupling. The observed differences in the effect of aCRT and conventional CRT on interventricular dyssynchrony suggest that a reduction of RV uncoupling together with adaptive AV optimization are the mechanisms behind the previously reported clinical benefit of aCRT.^3,4,6-9^

Adaptive CRT can exert its physiologic effects via two mechanisms: one involving the effect of the fusion of LV pacing with intrinsic RV activation on electrical dyssynchrony, and the other via adaptive AV optimization. Moreover, there is a complex interaction between interventricular dyssynchrony and AV optimization.^31, 32^ Our RCT showed that aCRT reduced RV uncoupling by significantly changing VEU, thereby reducing interventricular dyssynchrony, a known critical predictor of CRT response.^33, 34^ In the previous studies, VEU was strongly associated with clinical outcomes.^26^ The magnitude of change in VEU has been shown to be the primary driver of acute hemodynamic CRT response.^35^

Our results support previous experimental and clinical studies^10-13^ indicating similar effects of LV and BiV pacing on LV intraventricular dyssynchrony. It should be noted that in our study, the protocol did not mandate the placement of the LV lead in the location corresponding to the latest activation.^36, 37^ It is known that the degree of interventricular dyssynchrony depends on the location of LV and RV pacing, and the complex interaction of pacing sites with the heterogeneous substrate in CRT recipients.^38^ Nevertheless, the aCRT-ELSYNC RCT results suggest that both reduction of RV uncoupling and adaptive AV optimization are responsible for the previously reported advantages of aCRT over conventional CRT: higher likelihood of CRT response,^5^ and reduction of AF occurrence.^6-9^ Our results are consistent with previous observations concluding that an optimal AV delay is necessary for effective fusion of LV pacing with intrinsic excitation and the most advantageous reduction of interventricular dyssynchrony.^38^

In our study population, only a few participants met strict LBBB criteria, and nearly half of the participants had an S wave in V_5_-V_6_, which explains why we observed both RV- and LV-uncoupling.^26^ The presence of an S wave in V_5_-V_6_ has previously been associated with poor CRT response, HF re-hospitalizations and all-cause mortality.^39^ Bi-ventricular and RV enlargement^40^ and apical location of the LV posterior fascicular branch^41^ were suggested as possible mechanisms behind atypical LBBB with S wave in V_5_-V_6_. Our observation that conduction block in such atypical LBBB occurred in the anterior RV, as opposed to the interventricular groove in those who met the strict LBBB criteria, suggests yet another possible mechanism. Longitudinal dissociation of the His bundle and predestined conduction cables^42^ suggest that proximal conduction block of fibers that form downstream branches of both right and left bundles may produce atypical LBBB with S wave in V_5_-V_6_. We speculate that patients with this type of conduction abnormality should respond to His bundle pacing. This hypothesis should be tested in future prospective studies.

Similar to previous studies,^26, 43^ we observed considerable heterogeneity in epicardial activation sequences. As observed in previous studies,^35, 44, 45^ RV pacing and BiV pacing prolongs RVAT and increases RV uncoupling. In contrast, single-site LV pacing with fusion reduces RV uncoupling and interventricular dyssynchrony in a wide range of LBBB morphologies, including IVCD and atypical LBBB with S wave in V_5_-V_6_, as demonstrated in this RCT.

The aCRT-ELSYNC RCT results did not find a significant correlation between metrics of electrical dyssynchrony measured on a surface ECG and the epicardial surface; however, both sets of metrics showed a significant reduction in the degree of dyssynchrony six months post-CRT. This finding highlights the differences between the two approaches and their complementary value. The ECGI method^20^ utilized in this study is built on a pericardial potential source model, showing the distribution of electrical potentials on the pericardial surface of the heart. In contrast, surface ECG metrics of electrical dyssynchrony (QRS duration, QRS area^27^, and SAIQRST^14, 15, 25^) are global metrics of electrical dyssynchrony, a sum of dyssynchrony through endo-, mid-, epicardium, and both ventricles.^46, 47^ Through results from the epicardial activation maps metrics, we could appreciate the variation in LV activation patterns at baseline and the reduction in VEU in only the aCRT group. From the surface ECG, the QRS area and SAIQRST significantly decreased post-CRT in both groups. Using both metrics allows for a comprehensive evaluation of electrical dyssynchrony, with both showing an overall reduction in electrical dyssynchrony post-CRT.

A weak correlation between ECGI measures of electrical dyssynchrony and QRS duration has been previously reported.^26, 38, 48^ Similarly to many previous ECGI studies,^26, 30, 35, 43-45, 48^ we observed highly variable patterns of epicardial activation and response to LV and BiV pacing, highlighting the importance of individualized planning of both RV^49^ and LV lead placement and assessment of the pacing effect. Studies using an ECG belt showed that electrical dyssynchrony measured on the body surface could provide a good prediction of hemodynamic CRT response^50^ and LV remodeling,^51^ helping to optimize LV lead location. Simple VCG metrics (SAIQRST and QRS area) showed an advantage over QRS duration for the prediction of CRT response.^14, 15, 27^ Experimental studies showed limitations of ECGI-derived dyssynchrony indices, which had only a moderate agreement with directly measured interventricular dyssynchrony.^52^ Available resources might dictate the choice regarding method of assessment of electrical dyssynchrony (VCG, ECG belt, ECGI) in clinical practice. Further development of these methods of assessment of electrical dyssynchrony is essential to equip physicians in a wide range of practice settings.

### Limitations

The number of enrolled and randomized participants was modest. However, this is the first and largest RCT to date that studied detailed, noninvasively reconstructed epicardial mapping of electrical activation abnormalities in patients undergoing CRT. Leads placement was not standardized or guided by the study protocol; this requires further study. As we reconstructed activation on the epicardial surface only, the lack of information about septal and mid-myocardial activation limited assessment of the dyssynchrony by ECGI.

## Data Availability

Data can be made available to other investigators after confirmation of appropriate IRB oversight of the proposed additional analyses.

## Acknowledgments

The authors thank the study participants and staff. We thank William Woodward, ARMRIT, for the help with the CMR data acquisition.

## Supplemental Material

**Supplemental Table 1.** Correlation matrix for baseline (n=32) electrical dyssynchrony metrics measured on the reconstructed epicardial activation map and body surface ECG.

| | Spearman's $\rho$ coefficient | | | |
| --- | --- | --- | --- | --- |
|  | VEU | EDI<sub>V</sub> | EDI<sub>LV</sub> | EDI<sub>RV</sub> |
| QRS duration | 0.059 | 0.334 | 0.267 | 0.241 |
| *P-value* | *0.752* | *0.062* | *0.140* | *0.283* |
| QRS area | -0.047 | 0.091 | 0.261 | -0.114 |
| *P-value* | *0.797* | *0.622* | *0.150* | *0.536* |
| SAIQRST | 0.021 | 0.120 | 0.175 | 0.037 |
| *P-value* | *0.908* | *0.064* | *0.340* | *0.839* |

**Supplemental Figure 1.**
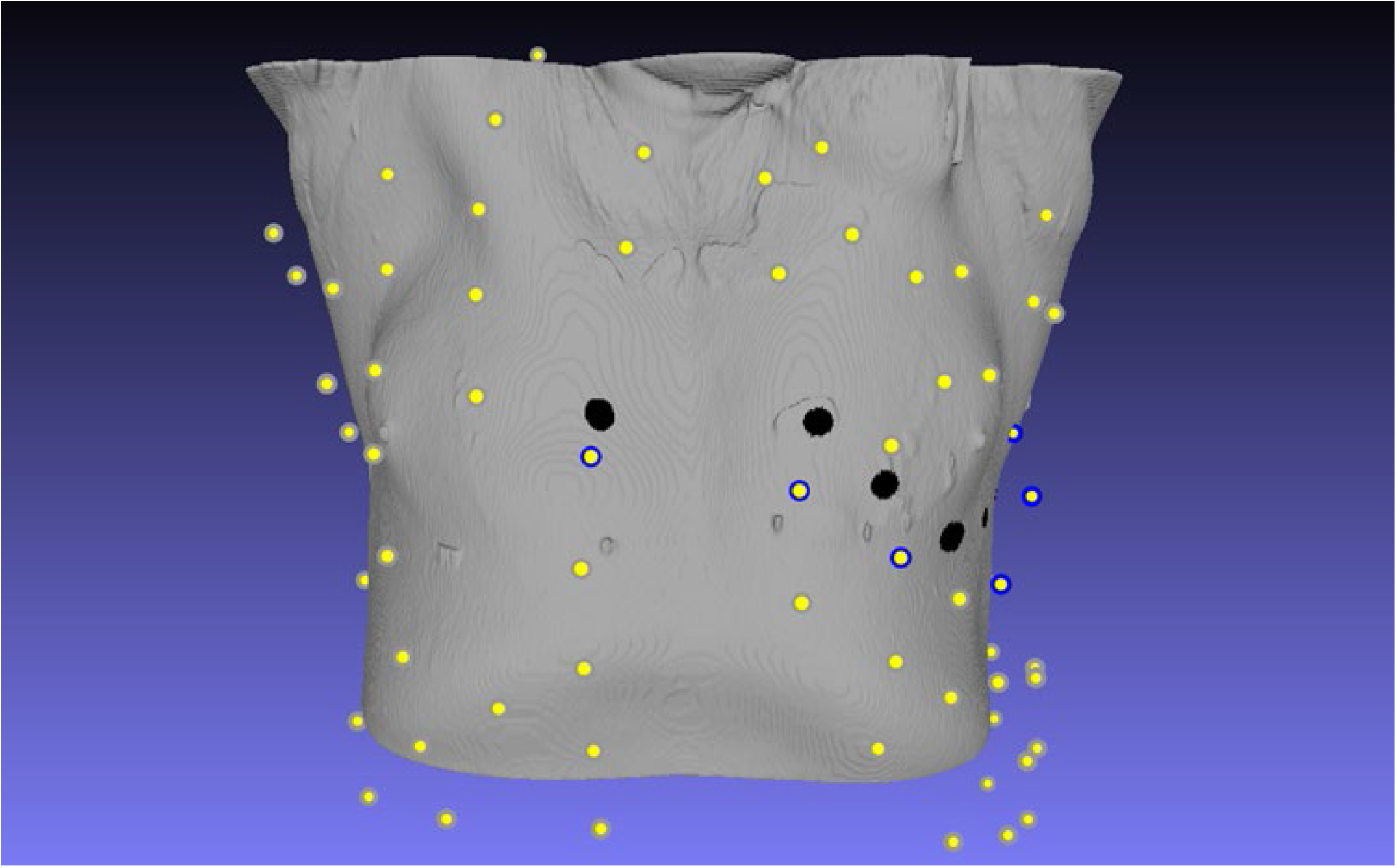
Approximation of standard 12 lead ECG location on from 128-lead electrode position. Black dots-Approximated location of precordial V1-V6 electrodes. Yellow dots – noise-free body surface mapping electrodes. Blue circled yellow dots indicated 6 nearest electrodes approximated the V1-V6 lead location of standard 12-lead ECG.

**Supplemental Figure 2.**
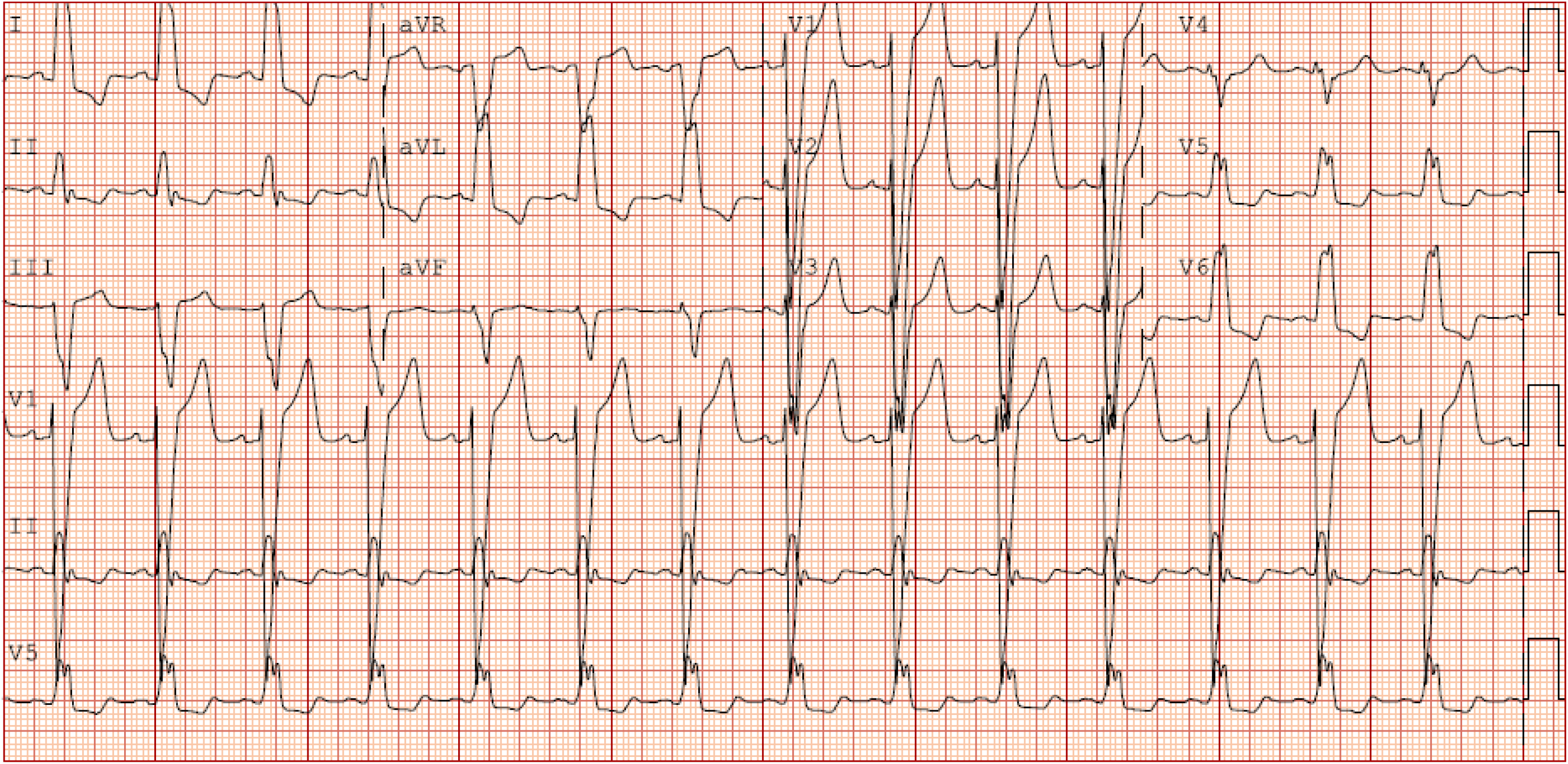
A representative example of a typical LBBB in a female study participant with nonischemic cardiomyopathy (the corresponding map is shown in Figure 3A).

**Supplemental Figure 3.**
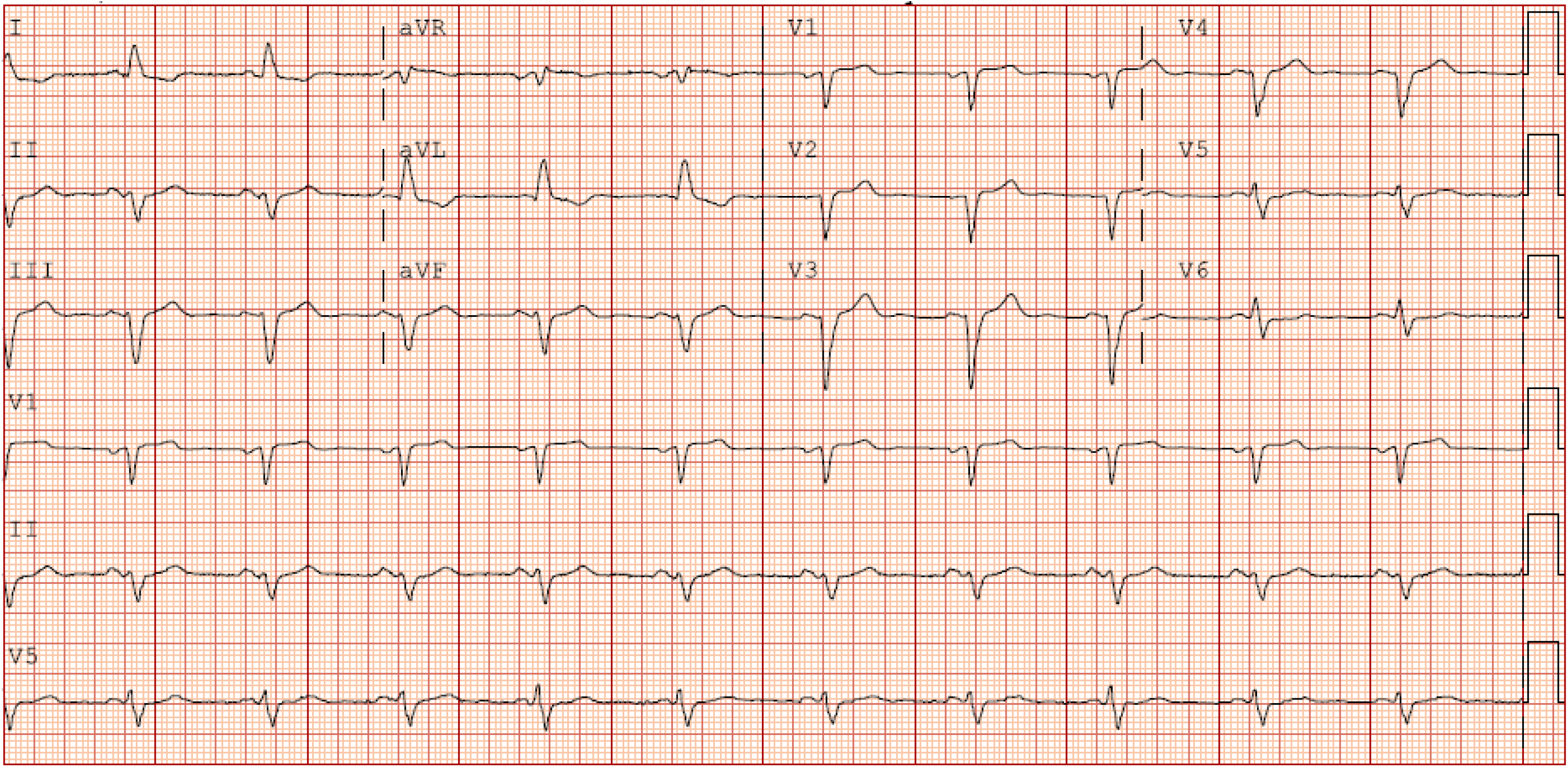
A representative example of interventricular conduction delay (atypical LBBB) in a female study participant with nonischemic cardiomyopathy (the corresponding map is shown in Figure 4A).

